# Microscale dynamics of electrophysiological markers of epilepsy

**DOI:** 10.1101/2020.10.14.20211649

**Authors:** Jimmy C. Yang, Angelique C. Paulk, Sang Heon Lee, Mehran Ganji, Daniel J. Soper, Pariya Salami, Daniel Cleary, Mirela Simon, Douglas Maus, Jong Woo Lee, Brian Nahed, Pamela Jones, Daniel P. Cahill, Garth Rees Cosgrove, Catherine J. Chu, Ziv Williams, Eric Halgren, Shadi Dayeh, Sydney S. Cash

## Abstract

**Objective:** Interictal discharges (IIDs) and high frequency oscillations (HFOs) are neurophysiologic biomarkers of epilepsy. In this study, we use custom poly(3,4-ethylenedioxythiophene) polystyrene sulfonate (PEDOT:PSS) microelectrodes to better understand their microscale dynamics.

**Methods:** Electrodes with spatial resolution down to 50µm were used to record intraoperatively in 30 subjects. For IIDs, putative spatiotemporal paths were generated by peak-tracking, followed by clustering. For HFOs, repeating patterns were elucidated by clustering similar time windows. Fast events, consistent with multi-unit activity (MUA), were covaried with either IIDs or HFOs.

**Results:** IIDs seen across the entire array were detected in 93% of subjects. Local IIDs, observed across <50% of the array, were seen in 53% of subjects. IIDs appeared to travel across the array in specific paths, and HFOs appeared in similar repeated spatial patterns. Finally, microseizure events were identified spanning 50-100µm. HFOs covaried with MUA, but not with IIDs.

**Conclusions:** Overall, these data suggest micro-domains of irritable cortex that form part of an underlying pathologic architecture that contributes to the seizure network.

**Significance:** Microelectrodes in cases of human epilepsy can reveal dynamics that are not seen by conventional electrocorticography and point to new possibilities for their use in the diagnosis and treatment of epilepsy.

**Highlights:**

- PEDOT:PSS microelectrodes with at least 50µm spatial resolution uniquely reveal spatiotemporal patterns of markers of epilepsy
- High spatiotemporal resolution allows interictal discharges to be tracked and reveal cortical domains involved in microseizures
- High frequency oscillations detected by microelectrodes demonstrate localized clustering on the cortical surface

## Introduction

Recent advances in microelectrode technology have revealed new detail of interictal discharges and seizures, including the appearance of events on the spatial order of microns. These microelectrode designs have captured data at high spatial resolution using penetrating or surface electrodes arranged in a grid (Schevon et al. 2008; Stead et al. 2010). Data from these microelectrode arrays have suggested that interictal and ictal activity can be generated from small areas on the order of 200 µm^2^, (Schevon et al. 2008) a finding that has in part been limited by electrode spatial resolution. Prior studies utilizing microelectrode arrays have relied on cortically invasive technologies, such as the NeuroPort array (Schevon et al. 2008; Keller et al. 2010) or, more often, have relied on electrode array designs with a minimum millimeter-level spatial resolution (Stead et al. 2010).

Progress in microelectrode technologies have led to increased flexibility in electrode arrangement and spatial resolution. Studies have reported on the use of organic electrode materials such as poly(3,4-ethylenedioxythiophene) polystyrene sulfonate (PEDOT:PSS) to record neural activity (Khodagholy et al. 2014, 2016), which can be lithographically patterned onto flexible parylene C substrates. These devices can be comprised of micron-level electrode contact sizes with high spatial resolution, while retaining low impedance (around 20-30 kΩ) and close surface conformability (Khodagholy et al. 2014). The combination of these unique properties is therefore hypothesized to allow for high signal-to-noise ratios, ultimately allowing for detection of single units from the cortical surface (Khodagholy et al. 2014, 2016).

Interictal discharges (IIDs) are neurophysiologic markers of epilepsy and may represent irritative cortex, though their cellular underpinnings and relationship to ultimate clinical outcome is debated (Wilke et al. 2009; Dworetzky and Reinsberger 2011). Nevertheless, they are considered neurophysiologic abnormalities that are taken into consideration during clinical management of patients who present with epilepsy (Wirrell 2010). More recently, high frequency oscillations (HFOs) have been identified as another marker for epileptic brain regions, though their role has been debated, as they can be considered either normal oscillations or pathological events (Worrell and Gotman 2011; Jefferys et al. 2012; Burnos et al. 2016; Cimbalnik et al. 2018). While definitions vary, HFOs have been separated into two sub-bands, the ripple band of 80-200 Hz and the fast ripple band of 250-500 Hz (Worrell and Gotman 2011). Early microelectrode studies argued that fast ripples may be increased in epileptogenic regions (Worrell and Gotman 2011; Thomschewski et al. 2019), and later clinical studies not only identified similar oscillations in macroelectrodes (Urrestarazu et al. 2007; Crépon et al. 2010) but also have suggested that resection of regions that generate fast ripples are correlated with seizure freedom (Fedele et al. 2017; Klooster et al. 2017). However, a more recent prospective study has suggested this may not be a consistent correlation (Jacobs et al. 2018). Finally, the intersection of these two markers, IIDs-ripples, have additionally been identified as potentially more specific markers for seizure onset zones (Klink et al. 2016), with their resection being correlated with improved post-surgical outcome (Wang et al. 2013, 2017).

We hypothesized that using higher spatial resolution PEDOT:PSS microelectrodes would not only reveal microdomains of interictal activity but also show particular dynamics that may not have been previously seen on microelectrodes with lower spatial resolution. In addition, we sought to demonstrate that these technologies could be adapted for acute intraoperative recordings as an exploratory measure to pave the way for future work involving microelectrodes with contacts on the order of thousands. We demonstrate that microelectrodes are not only able to show similar signals to clinical gold-standard electrocorticography electrodes but are also able to avoid spatial averaging effects and elicit information of highly localized processes with low noise. By revealing local IID and HFO events, as well as periodic discharges and seizures, we highlight that microelectrode technology offers insight into underlying epileptic processes that may be overlooked by current clinical technologies. Ultimately, our multi-faceted analyses support the hypothesis that local microdomains can demonstrate epileptiform activity.

## Methods

### Subjects

Recordings were performed in subjects who were already scheduled for a neurosurgical procedure at Massachusetts General Hospital (MGH) or Brigham and Women’s Hospital (BWH). This study was approved by the Partners Institutional Review Board, which covers both MGH and BWH. All subjects voluntarily participated and provided informed consent.

### Device Manufacture

The fabrication of the PEDOT:PSS device is similar to previously established protocols and are described elsewhere(Sessolo et al. 2013; Ganji et al. 2018). In this study, two different electrode designs were used. One was organized as a bi-linear array, with 128 channels arranged in two columns (each column with 64 channels), with an electrode diameter of 30 µm and interelectrode distance of 50 µm. A second design was comprised of a circular grid, arranged as concentric rings at varying distances from the center (Figure 1A). The use of one or the other electrode type was to test whether the bi-linear array (the first design used) or the circular grid could be better optimized for sampling IIDs and HFOs. The rationale for a bi-linear array was to allow for a maximum length of cortex to be investigated, in case specific boundaries of microdomains could be detected. The circular grid was arranged with gradually increasing distances between rings in order to maximize spatial coverage. Both electrode types appeared to detect our targeted events with the circular grid enabling a wide sampling of the signal at different spatial scales.

**Figure 1.**
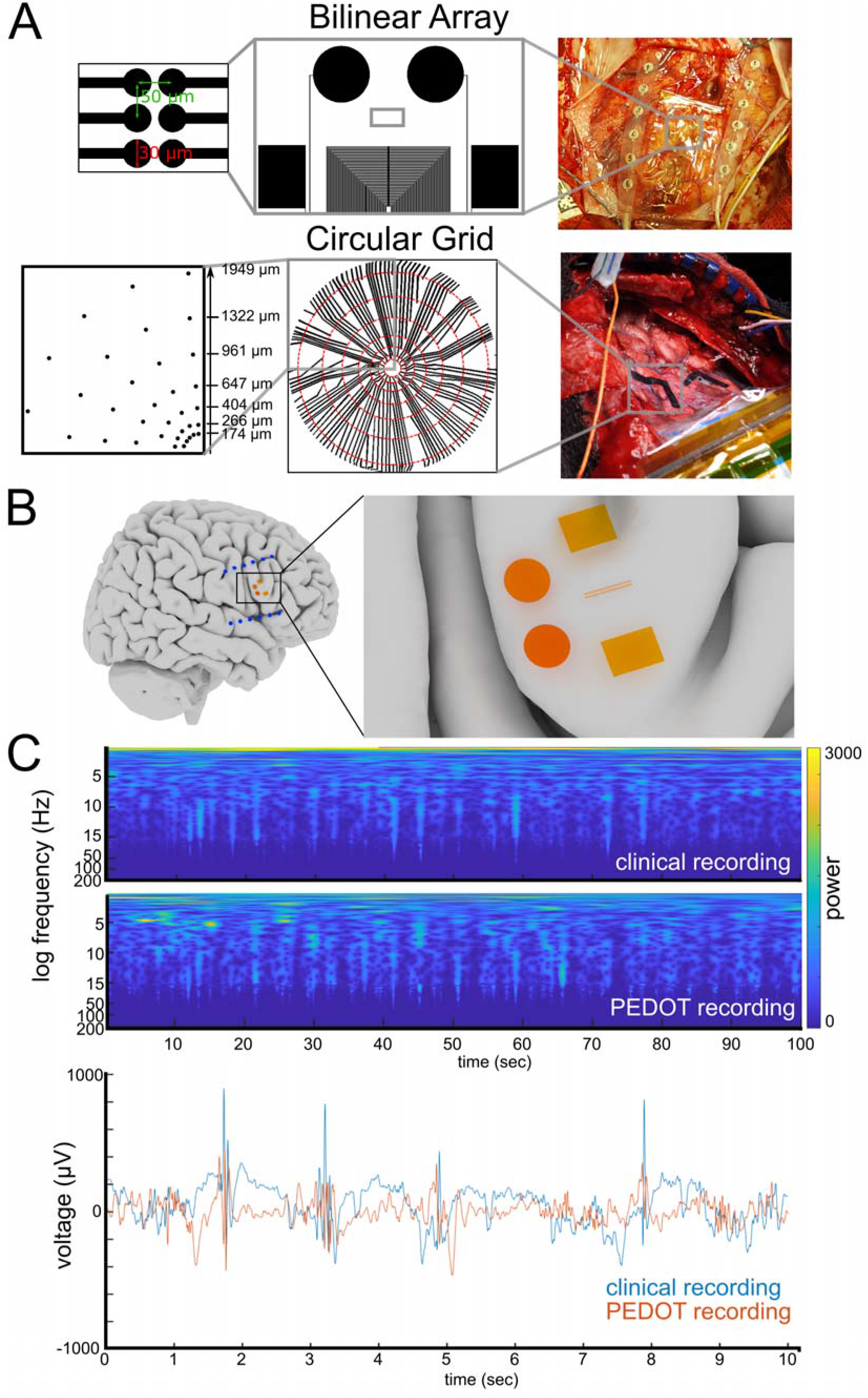
a) Two designs of PEDOT:PSS electrodes used, with intraoperative images. Top row shows the “bilinear array” comprised of two rows of 64 30µm-contacts, with 50µm spacing. The entire length of the electrode is 3150µm (center-to-center). Bottom row shows the “circular grid,” comprised of seven rings, with varying distances from the center. Intraoperative photograph shows two circular grids on the cortical surface. b) 3-dimensional reconstruction of subject’s brain, with overlaid putative positions of clinical strip electrodes and PEDOT:PSS bilinear array. The bilinear array is the two small lines in the expanded left panel. The two circles and the two squares are localizing markers to indicate the placement of the electrode and allow for photographic alignment of the recordings in the case (see (a)) c) Comparison of clinical recording vs PEDOT:PSS experimental recording. LFP (Top: clinical recording, Bottom: PEDOT:PSS recording) and raw voltage traces (Blue: clinical recording, Red: PEDOT recording) are demonstrated.

### Data Acquisition and Processing

Research recordings were conducted using an Intan recording system with a sampling rate of 30 kHz (filtered from 1 Hz to 7500 Hz to reduce aliasing) as described previously(Hermiz et al. 2016). During acquisition, ground and reference needle electrodes (Medtronic, Minneapolis, MN, USA) were placed in nearby tissue, normally in muscle or scalp. After electrode placement, impedance testing was performed using the Intan RHD2000 software. OpenEphys, an open-source electrophysiology software suite, allowed for recording and visualization across all channels along with analog triggers to enable synchronization of the signals (Siegle et al. 2017).

When used, clinical electrocorticography or depth electrode recording was performed simultaneously using PMT or Ad-tech electrodes (Ad-tech Medical, Racine, WI, USA, or PMT, Chanhassen, MN, USA). In general, at BWH, Ad-tech clinical strip platinum electrodes utilized 10 mm spacing with 2.3 mm contact diameter, and Ad-tech depth platinum electrodes had 5-8 mm spacing with a 2.41 mm contact size, with a 1.12 mm diameter. In general, at MGH, PMT Cortac clinical strip platinum electrodes had 10 mm spacing with 3 mm contact diameter, and PMT Depthalon depth platinum electrodes had 3.5 mm spacing with 2 mm contacts with a 0.8 mm diameter. Clinical electrodes were placed by the attending neurosurgeon in the regions of clinical interest. In general, the Natus Quantum system (Natus Neurology Inc., Middleton, WI, USA) was used for clinical recordings, with sampling rates as dictated by the clinical team and typically at either 4096 or 512 Hz.

### Data Analysis

Data analysis was performed using custom MATLAB scripts. Raw voltage signals were taken directly from the Intan recordings. For interictal discharge (IID) detection and analysis, local field potential (LFP) data were decimated to 1000 Hz, demeaned relative to the entire recording, and line noise and its harmonics up to 200 Hz were removed by subtracting the band-passed filtered signals from the raw signal. For detection of high frequency oscillations (HFOs), raw data were decimated to 2000 Hz and band-passed in a frequency range of 80-200 Hz for the ripple band and 250-500 Hz for the fast ripple band as previously reported (Lévesque et al. 2012; Salami et al. 2012). Due to the potential for noise in an intraoperative environment which would alter the analyses, recordings and channels were scrutinized for noise. Channels with excessive line noise, artifacts, or high impedances (>100 kΩ) were removed from the analysis. The recordings of 3 subjects were removed due to movement artifacts during a majority of the recording. In examining the high frequency domain for the analysis of HFOs, an additional 3 recordings were removed due to high frequency electrical noise, likely due to the intraoperative environment, that could not be suppressed.

Interictal discharges were automatically detected using an algorithm that first filters the data into a 10-60 Hz band, applies an envelope and then finds an appropriate threshold value modeled on a statistical distribution in the sampled envelope (Janca et al. 2015). Each detection was subsequently reviewed visually. IIDs were classified into either a general event, in which it was seen across >50% of channels, or a local event, which was seen across ≤50% of channels. For recordings in which at least 10 IIDs were found, peak tracking was performed for the interictal discharge on each channel by finding the maximum or minimum value for each interictal discharge event, which provided a path for that event across electrode sites over a specified time window of 2 seconds. While long, considering the propagation of the path, it ensured that we did not clip any possible path information. The discrete Fréchet distance between pairs of events were subsequently calculated to determine path similarity (Eiter and Mannila 1994). In brief, Fréchet distance is a measure of path similarity by calculating, and minimizing, the distance between them. This script represents an approximation by using sampled points along each curve. Since the path similarity was measured across the same electrode array per recording (and not compared across recordings), the Fréchet distance measured allowed a within-array comparison across IIDs. In addition, as the Fréchet distance measure is a point-by-point comparison between paths, it is not as susceptible to the differences in electrode spacing or distributions as it is a distance measure between paths on a point-by-point basis. These values were subsequently clustered by evaluating solutions as provided by a MATLAB agglomerative clustering algorithm, using a silhouette clustering evaluation criterion. The mean coordinate for each discharge, per clustered group, was then calculated, and a smoothed mean over 10 ms was applied in order to find the average trajectory of that clustered group.

HFOs were automatically detected using a previously published algorithm (Lévesque et al. 2012; Salami et al. 2012). In brief, the detector first filters the data into two bands, a ripple band (80-200 Hz) and a fast ripple band (250-500 Hz). It subsequently uses a reference time window in order to detect peaks in the filtered data. A series of at least 4 oscillations was necessary to be considered an HFO. To further avoid artifacts and false detections, only fast ripples that did not overlap with the lower frequency ripples were considered detections. Each channel was additionally visually reviewed manually for thresholding to validate and verify the oscillatory waveforms as well as to check the raw recordings to ensure that the oscillations were not due to sharp large voltage events. Overall, detections were at least 2.9 standard deviations above the mean when addressing amplitude criteria for the detector, in terms of thresholding for determining true HFOs. The morphology of HFOs was not distinctly considered given prior research suggesting that this attribute does not distinctly improve delineation of epileptogenic zones (Burnos et al. 2016).

To determine spatial similarity of detections of HFO events over the electrode array throughout the recording, an approach that has been previously described to analyze the neural patterns known as avalanches was adapted (Beggs and Plenz 2004; Ribeiro et al. 2016). In brief, the average inter-event time was determined, and then, to avoid excessive simplification of patterns, the time window was increased by 5 standard deviations. A similarity index was then calculated between time windows as defined by Sim(A,B) = |A ∩ B| / |A ∪ B|. These similarity indices between time frames were then clustered using a paired clustering algorithm, based on Euclidean distance, leading to ‘families’ of time-frames that had optimal similarity. The significance of each family was then compared to 1,000 shuffled datasets that were created by randomly permuting the detections per channel in all time frames, which allowed the overall number of active electrodes in each time window to remain constant. The probability for obtaining a family of a given size and average similarity was then calculated using the same clustering method as for the actual data, and the Benjamini-Hochberg multiple comparisons correction was implemented (Ribeiro et al. 2016).

Fast unitary waveform events (similar to single or multi-unit activity) were detected and sorted into clusters of waveforms using Kilosort (Pachitariu et al. 2016). Kilosort detects high frequency waveforms and then clusters these waveforms based on both the waveform shape as well as the spatial mapping of the waveforms on the electrode grid. We were able to identify repeated waveform events which could be clustered similar to typical single unit spike. To eliminate artifact, we only applied Kilosort to data which had no movement or stimulation artifact or we removed noisy channels to detect clear putative clusters of waveform events.

Further steps included rejection of clusters as sampled which looked to be artifact (such as with stimulation) or inter-event intervals (IEIs) corresponding with 60 Hz noise or IEIs which were too short (<2ms). We also identified the waveforms in the raw and original high pass filtered data to confirm their timing and channel location. We then pooled the waveform times and identified these pooled times as fast events similar to multi-unit activity (MUA). The reason we did not solely threshold the activity and detect events from the recordings was to be conservative in these measures and to ensure we were measuring what could be MUA activity while eliminating any possible artifacts.

To understand if a relationship existed among interictal discharges, high frequency oscillations, and fast events, covariance between two groups of detections was calculated for each data set. Mean values of HFO covariance either 200ms before or after the interictal discharge were found, and the non-parametric test, Wilcoxon Rank Sum, was used to calculate whether the central peak was significantly different from a 1 second period, 5 seconds before the peak across subjects. For comparisons with fast events, values of covariance either 5 seconds before or after the interictal discharge or HFO were found, and the non-parametric test, Wilcoxon Rank Sum, was used to calculate whether the central peak was significantly different from a 1 second period, 5 second before the peak across subjects. This comparison was done on a per-time point level (every 0.05 sec) and corrected for multiple comparisons using a false discovery rate control.

## Results

### Subjects

In this work, 30 subjects participated, with a mean age of 39.5 (standard deviation of 12.5) with 14 men and 16 women (Table 1). A bi-linear electrode array, as opposed to a circular array, was used in 63% of the subjects (Figure 1A), with the electrode typically situated on an exposed gyrus within the surgical field (Figure 1B). The majority, 83%, of subjects were right-handed, and 73% underwent a neurosurgical procedure for epilepsy. The remaining patients underwent a neurosurgical procedure for either tumor resection or vascular malformation resection. As these patients can also present with seizures, 93% of subjects overall had prior seizures. General anesthesia was used in 67% of the subjects. Of the patients undergoing a neurosurgical procedure for epilepsy, 9 (41%) were treated with a medication, as part of their intraoperative clinical management, to activate epileptiform abnormalities. A comparison with simultaneous clinical electrocorticography recordings demonstrated the accuracy of the PEDOT:PSS recordings, with unique features such as IIDs appearing simultaneously in both recordings, demonstrating that similar neurophysiologic features could be detected using our experimental PEDOT:PSS microelectrode (Figure 1C).

**Table 1:** Subject and recording characteristics.

| Patient Number | Age | Handedness | Type of Case | Anesthesia Type | Type of Electrode | Inducing Medication | Cold Saline | Recording Length (Seconds) | Interictal Discharges |  |  |  | HFOs Number of Clusters |
| --- | --- | --- | --- | --- | --- | --- | --- | --- | --- | --- | --- | --- | --- |
|  |  |  |  |  |  |  |  |  | General IIDs | Local IIDs | General IID Paths | Local IID Paths |  |
| 1 | 30s | Right | Left temporal lobectomy for seizure | General | Linear | Methohexital |  | 1250 | 32 | 0 | 3 |  | 12 |
| 2 | 30s | Right | Right temporal lobectomy for seizure | General | Linear | Methohexital |  | 628 |  |  |  |  |  |
| 3 | 20s | Right | Right temporal lobectomy for seizure | General | Linear | Methohexital |  | 717 |  |  |  |  |  |
| 4 | 20s | Right | Right temporal lobectomy for seizure | General | Linear |  |  | 1213 | 2 | 2 |  |  | 5 |
| 5 | 40s | Left | Right temporal lobectomy for seizure | General | Linear | Methohexital |  | 941 | 118 | 2 | 8 |  | 31 |
| 6 | 20s | Right | Right temporal lobectomy for seizure | General | Linear | Methohexital |  | 1377 | 50 | 0 | 5 |  |  |
| 7 | 40s | Right | Right redo temporal lobectomy for seizure | General | Linear | Methohexital |  | 1501.5 | 167 | 2 | 2 |  | 27 |
| 8 | 30s | Right | Right temporal lobectomy for seizure | General | Circular | Methohexital | Cold saline | 2205 | 49 | 30 | 2 | 2 | 17 |
| 9 | 30s | Right | Left parietal tumor resection | MAC | Linear |  |  | 445 | 2 | 1 |  |  | 6 |
| 10 | 40s | Right | Right temporal lobectomy for seizure | General | Linear |  | Cold saline | 544.5 |  |  |  |  |  |
| 11 | 40s | Right | Right temporal lobectomy for seizure | General | Linear | Alfentanil |  | 1003 | 48 | 0 | 7 |  |  |
| 12 | 20s | Left | Right redo parietal resection for seizure | General | Linear |  |  | 348 | 0 | 0 |  |  | 14 |
| 13 | 60s | Right | Left temporal tumor resection | MAC | Linear |  |  | 715 | 1 | 7 |  |  | 24 |
| 14 | 30s | Right | Right parietal tumor resection | General | Linear |  |  | 301 | 4 | 1 |  |  |  |
| 15 | 30s | Right | Left redo temporo-parietal resection for seizure | MAC | Linear |  | Cold saline | 346 | 2 | 0 |  |  | 5 |
| 16 | 20s | Right | Right temporal lobectomy for seizure | General | Linear |  |  | 844.5 | 9 | 0 |  |  | 2 |
| 17 | 50s | Right | Left temporal craniotomy for AVM | MAC | Linear |  |  | 887.5 | 1 | 5 |  |  | 16 |
| 18 | 20s | Left | Right redo parietal resection for seizure | General | Circular |  |  | 1191 | 108 | 35 | 4 | 2 | 2 |
| 19 | 50s | Left | Right temporal tumor resection | MAC | Circular |  | Cold saline | 961 | 7 | 22 |  | 2 | 8 |
| 20 | 40s | Right | Right temporal lobectomy for seizure | General | Circular |  |  | 546.5 | 13 | 2 | 4 |  | 1 |
| 21 | 30s | Right | Right frontal lobectomy for seizure | General | Circular |  | Cold saline | 1901.5 | 136 | 6 | 28 |  | 1 |
| 22 | 50s | Right | Left craniotomy for tumor | MAC | Linear |  | Cold saline | 600 | 193 | 47 | 30 | 9 | 15 |
|  |  |  |  |  |  |  |  | 785 | 163 | 45 | 11 | 6 | 51 |
| 23 | 30s | Right | Left craniotomy for tumor | MAC | Circular |  | Cold saline | 785 | 1 | 0 |  |  | 9 |
| 25 | 50s | Right | Right temporal lobectomy for seizure | General | Circular |  | Cold saline | 1370.5 | 29 | 8 | 8 |  | 1 |
| 26 | 50s | Right | Right temporal lobectomy for seizure | General | Circular | Alfentanil |  | 975.6 | 1 | 0 |  |  | 0 |
| 27 | 20s | Left | Left modified hemispherectomy | General | Circular |  |  | 377.6 | 0 | 0 |  |  | 0 |
|  |  |  |  |  |  |  |  | 941.8 | 3 | 0 |  |  | 0 |
| 28 | 20s | Right | Right temporal lobectomy for seizure | General | Circular |  |  | 798 | 4 | 0 |  |  | 0 |
| 29 | 60s | Right | Left craniotomy for tumor | MAC | Linear |  |  | 945.2 | 11 | 0 | 5 |  | 2 |
| 30 | 50s | Right | Left temporal lobectomy for seizure | MAC | Linear |  |  | 106 | 1 | 0 |  |  | 0 |
|  |  |  |  |  |  |  |  | 1760 | 27 | 4 | 9 |  | 3 |
|  |  |  |  |  |  |  |  | 521.2 | 9 | 3 |  |  | 0 |
| 31 | 30s | Right | Left frontal craniotomy for seizure | MAC | Circular |  |  | 1358.3 | 69 | 13 | 13 | 3 | 0 |
|  |  |  |  |  |  |  |  | 687.7 | 25 | 5 | 3 |  | 5 |

### Interictal discharges

General interictal discharges were defined as events that, upon visual review of automated detections, were seen over more than 50% of the electrode array (Figure 2A), whereas local events were seen on fewer than 50% of the channels (Figure 2B). General IIDs were seen in 93% of the subjects, whereas local IIDs were less frequent and only seen in 53% of the subjects. The minimum distance required to detect a local IID was found to be 100µm. In examining the responses of interictal discharges to intervening measures, we found that the use of activating medications significantly increased the frequency of general interictal discharges by 180 seconds after administration, and the use of cold saline irrigation decreased the frequency of general interictal discharges by 240 seconds after the start of irrigation (Figure 2C, 2D). There was no statistically significant change in local interictal discharges with these intervening maneuvers.

**Figure 2:**
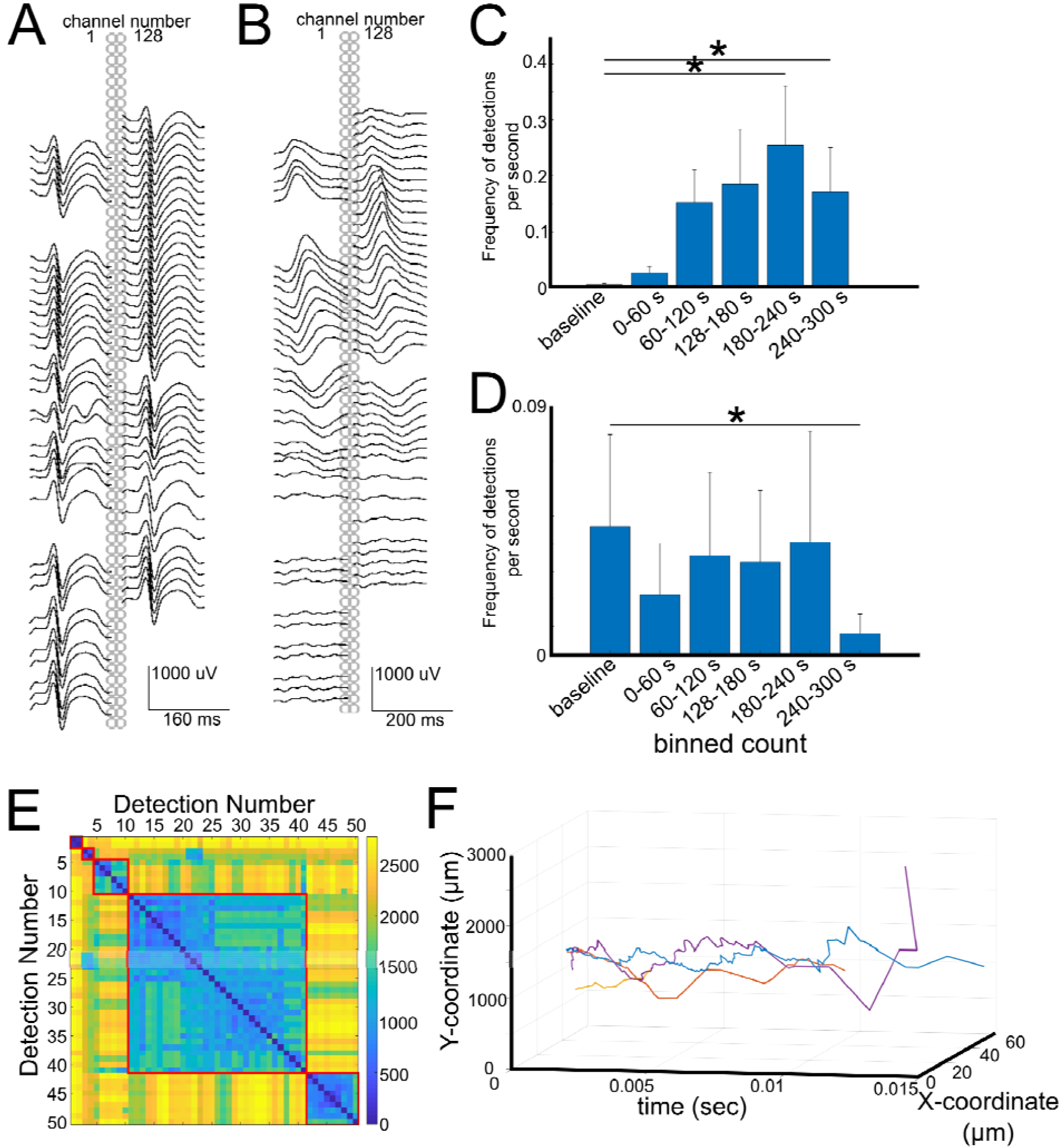
Interictal Discharges. a) Example of a general interictal discharge that was seen over the entire PEDOT:PSS bilinear array, with low pass filtering as described in Methods. Electrode number 1 and 128 are indicated. b) Example of a local interictal discharge that was seen only over a portion of the PEDOT:PSS bilinear array, organized in the same way as (a). c) Response of interictal discharges to activating medications (methohexital or alfentanil), N=7. * indicates p<0.05 (Wilcoxon Rank-Sum). d) Response of interictal discharges to cold saline irrigation, N=7. * indicates p<0.05 (Wilcoxon Rank-Sum). e) Example of clustering by Fréchet distance. f) Example of clustered families, and mean vector for each family represented in 3- dimensional space.

In order to decipher whether a phenomenon of “traveling” could be seen with interictal discharges, in which electrode contacts detected the same interictal discharge at different time points, the maximum or minimum voltage of the interictal discharge was pinpointed around a 2 second time window. To understand whether these “paths” could be grouped into specific patterns, the discrete Fréchet distance was calculated and values were clustered (Figure 2E). This approach revealed that at least 2 paths could be clustered across IIDs for each subject when examining general IIDs (Figure 2F). The overall average speed of these interictal discharges was found to be 60 ± 3.5 mm/s (standard error of the mean, SEM).

### High Frequency Oscillations

High frequency oscillations were seen in all recordings. Overall, high frequency oscillations also appeared to be responsive to manipulating measures. The use of medications that are known to promote the appearance of interictal discharges (alfentanil or methohexital) resulted in an increase in the average frequency per channel of HFO detections, but this change was not statistically significant (Figure 3A). On the other hand, the frequency of these detections decreased significantly 60 seconds after the initiation of cold saline irrigation (p<0.05, Wilcoxon rank-sum).

**Figure 3:**
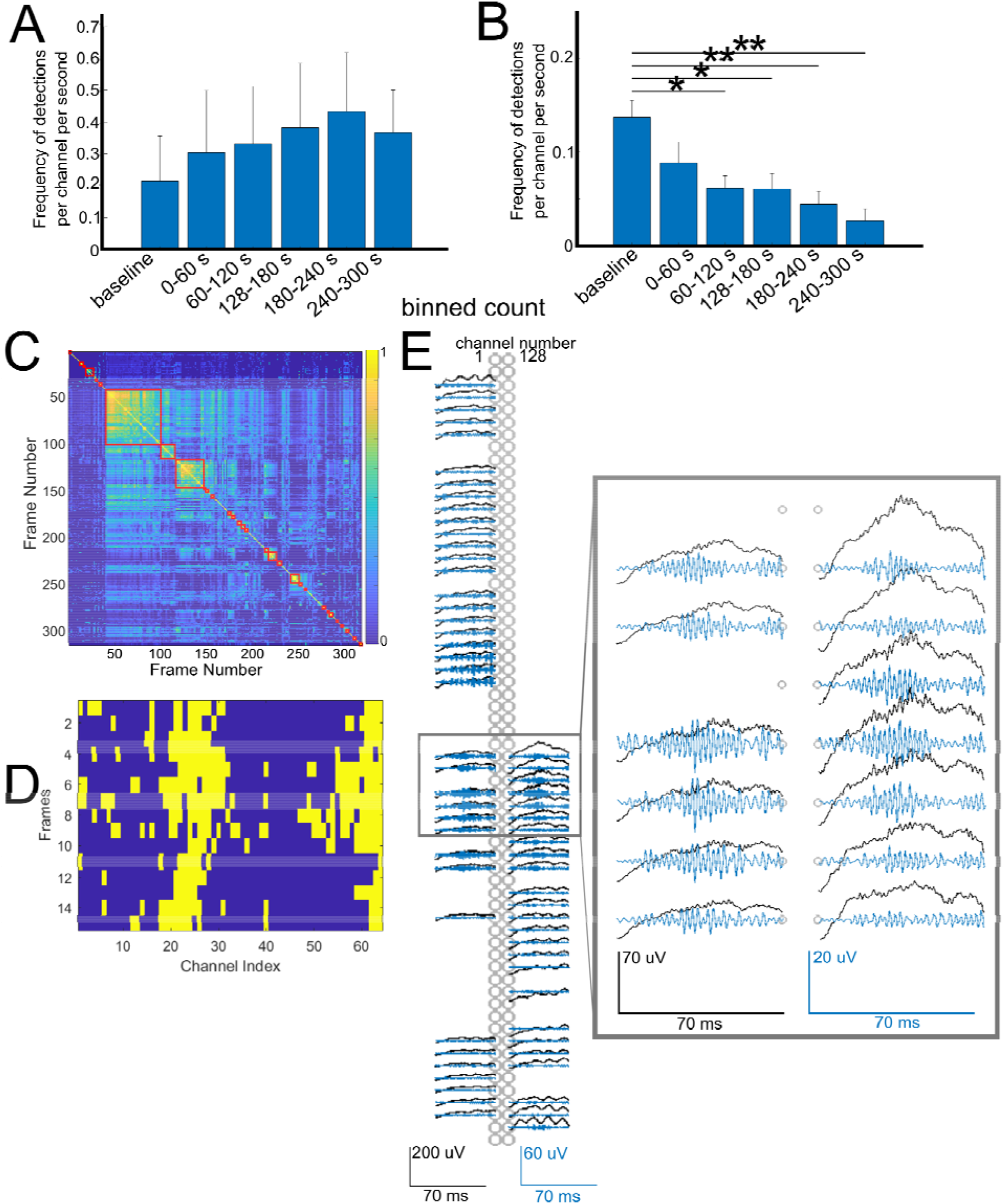
High Frequency Oscillations. a) Response of average frequency of HFO detections per channel to activating medications (alfentanil, methohexital). b) Response of average frequency of HFO detections per channel to cold saline irrigation. * indicates p<0.05, ** indicates p<0.01, Wilcoxon rank-sum. c) Example of similarity-based clustering by frames in one subject. d) Example of one group of clustered frames from (C). e) Demonstration one member of the group seen in (D), with black lines demonstrating low pass filtered data and blue lines demonstrating the fast ripple filtered band. Inset demonstrates a portion of the array in magnified view.

Analysis of the HFOs across channels demonstrated that a single HFO could appear in a subset of channels at the same time. In examining the patterns of HFO detections across channels in time windows, it was found that a similar pattern of detections in particular channels could be clustered (Figure 3C) into separate groups spatially among subsets of channels (Figure 3D, Figure 3E). By using a threshold of p≤0.001, 88% of subjects demonstrated at least 1 pattern of HFO detections that was unique and could not be explained by chance (as compared with a shuffled data set). At minimum, one electrode contact was involved in a significant family group.

Calculation of cross-covariance between interictal discharges and high frequency oscillations demonstrated whether a temporal relationship could be elucidated. For both general and local interictal discharges, it was found that HFO detections significantly preceded the interictal discharge using a 0.2s time window around the IID peak time (p<0.01, Wilcoxon rank-sum).

### Periodic Patterns and Microseizures

Rare examples were found of repeating interictal discharges, which are generally described as periodic discharges in a clinical setting. Two subjects demonstrated convincing representations of these localized patterns in few electrodes (Figure 4A, 4B). Given the lack of significant evolution or spread, these patterns were ultimately considered to be periodic patterns rather than distinctly seizure events.

**Figure 4:**
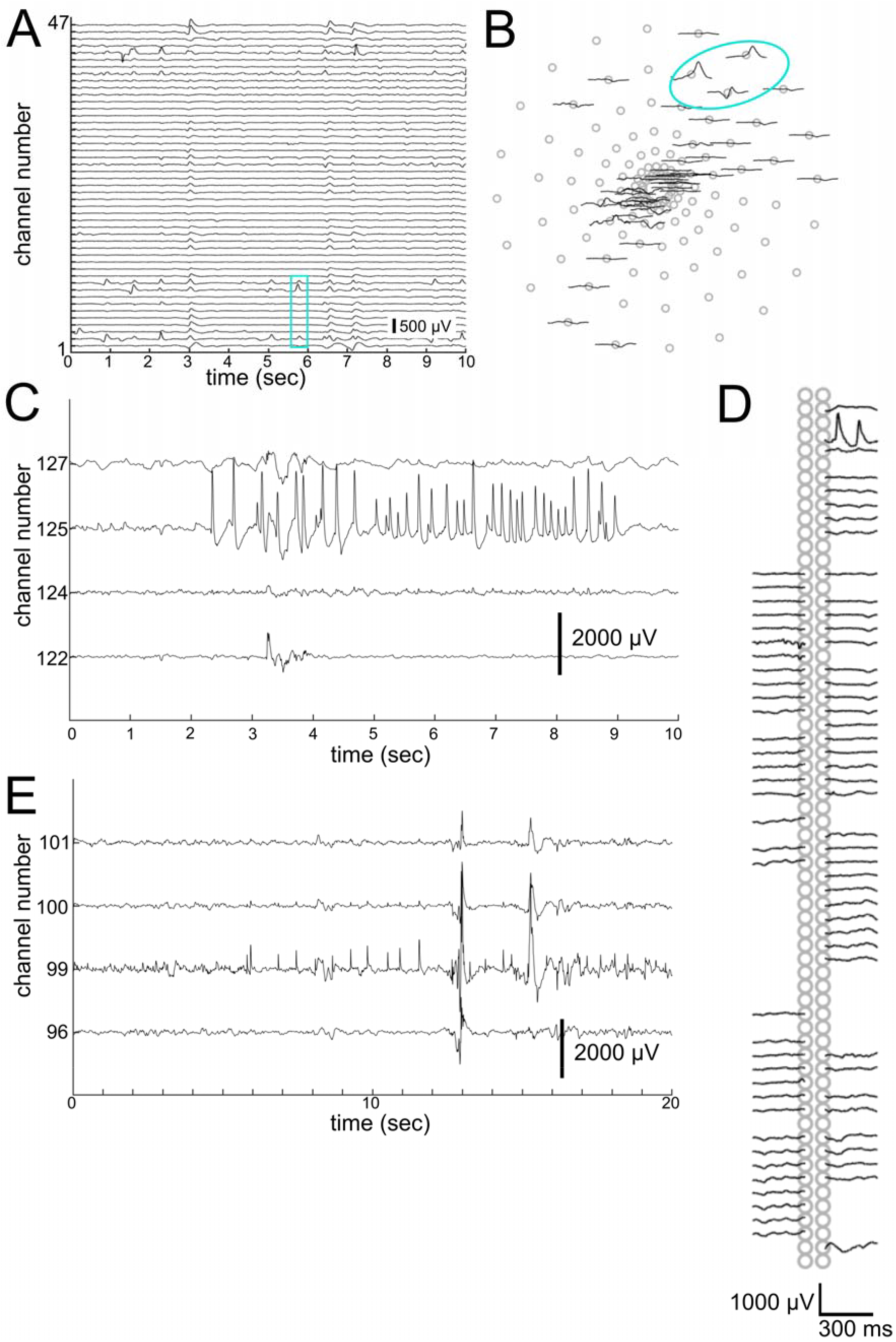
Periodic discharges and Microseizure events in PEDOT:PSS Microelectrodes. a) Demonstration of one series of periodic discharges, seen primarily in channels 6, 13, and 14. Low pass filtered data is shown, with distance between channels corresponding to 250µV. b) Spatial location of one of the series of periodic discharges as indicated in cyan box in A. c) One example of microseizure event, localized to channel 125. Field can be seen to extend to neighboring channels in 124 and 127. d) One discharge from C demonstrated spatially on the bi-linear array. e) Second example of microseizure event, localized to channel 99. Field can be seen to extend to neighboring channels 100. Distance between channels corresponds to 2000µV.

Examples of microseizure events were also found in one subject, who had history of seizures, which were isolated to primarily one electrode with a surrounding field (Figure 4C, 4E). These patterns were found to have highest amplitude in one electrode and distinctly demonstrated evolution in the frequency of discharges, which appeared to be consistent with a seizure-type phenomenon. Voltage fields to neighboring electrodes 50µm away were observed, though the microseizure event signal was highly localized. As the patient was under monitored anesthesia care (MAC), no clinical seizures were noted intraoperatively.

### Fast Events

We used the Kilosort software package (Pachitariu et al. 2016) to sort fast (>250 Hz) unitary events into a number of separable clusters using a template matching approach that considers waveform shape and waveform spatial spread across electrode sites (Figure 5). We arrived at clusters of detected repeated unitary fast events in 16 out of 30 participants that resembled multi-unit activity (MUA), which were pooled, and appeared similar to prior reports (Khodagholy et al. 2014). We could find these fast events colocalized with the general IIDs as well as with HFOs both in space (on the same electrode sites) and through time in single recordings (Figure 5A-C).

**Figure 5:**
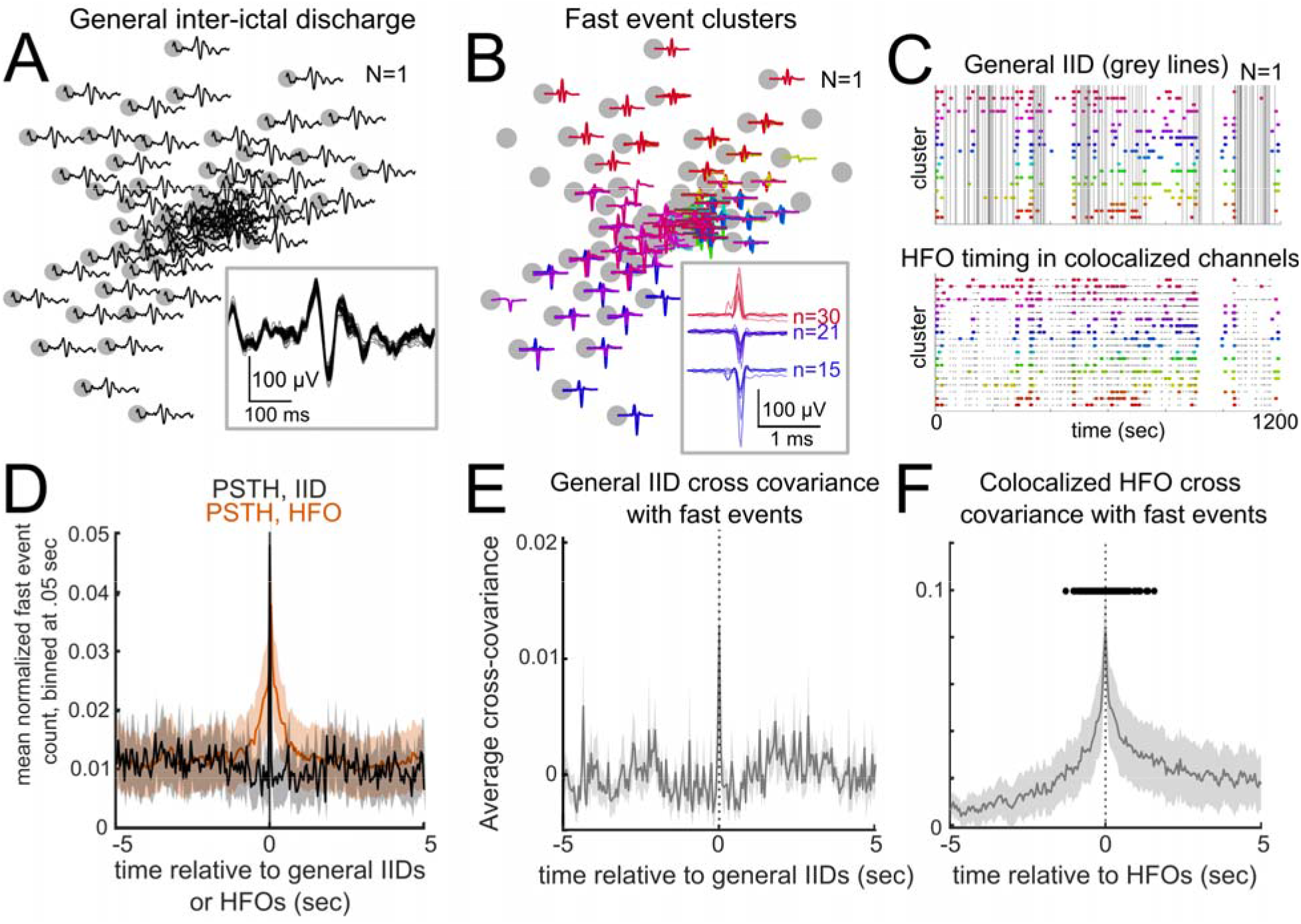
Multi-Unit Activity (MUA) a) Example spatial spread of a general IID across a circular grid (LFP filtered at <1000 Hz). Inset: zoomed in example of the IID with the waveforms from across the array overlaid on one another. b) Sorted clusters of fast (>250 Hz) unitary waveforms as distributed across the same circular grid array as shown in a). Different color lines are different clusters of unitary events. Inset: example waveforms for a few of the clusters. c) Raster plot showing the same recording in a) and b) showing the timing of the fast events in b) throughout the recording alongside the simultaneous general IIDs (grey lines in the top plot) and detected HFOs (grey dots in the bottom plot) in the same recording. d) The fast events were sorted using Kilosort (see Methods) and then pooled per recording similar to multi-unit activity (MUA) and then were compared with the timing of the general IIDs (black line) and HFOs (orange line) in peri-stimulus time histograms (PSTH). Lines are averages across recordings (after averaging PSTHs per recording). Shaded areas indicate standard error. e) The pooled fast event times were then covaried with the general IIDs per recording. The grey line is an average across recordings (after averaging cross-covariance curves per recording). Shaded areas indicate standard error. N=16. f) The pooled fast event times were then covaried with detected HFOs per recording. Black dots indicate the cross-covariance values are significantly different to zero. (Wilcoxon rank-sum test, p<0.05, false discovery rate controlled). The grey line is an average across recordings (after averaging cross-covariance curves per recording). Shaded areas indicate standard error. N=16.

To identify whether there was a temporal relationship between these aggregate fast events and the IIDs and HFOs, we calculated the peri-stimulus time histogram of the MUA fast events relative to the IIDs and HFOs (Figure 5D). We found peaks in the PSTH values at around zero, particularly for the HFOs, so there were some fast events which were occurring around the same time as the general IIDs and HFOs. To test this further, we calculated the cross-covariance values between the general IID times and the fast event times (Figure 5E) and the HFOs and the fast events (Figure 5F) across the data set. To determine if there is a significant cross-covariance relationship that is consistent across the data set between IIDs or HFOs and MUA, we calculated whether the cross-covariance values are significantly different to periods of time before the event (Figure 5E-F). We found the peaks in the cross-covariance averages between the general IIDs and the fast events were not significant relative to 5 seconds before the IIDs (Figure 5E). In contrast, we found the central peak in the cross-covariance averages between the HFOs and the fast events were significant relative to 5 seconds before HFOs (p<0.05, Wilcoxon rank-sum test, false discovery rate controlled, Figure 5E). This covariance relationship was not found between local IIDs and fast events (MUA) in either the PSTH or the cross-covariance calculations (data not shown). In other words, MUA co-varies, and could be occurring at the same times, as HFOs in these high spatial and temporal resolution recordings, but we could not find a covarying or time-locked relationship between IIDs and MUA in our recordings across patients.

## Discussion

We found that high spatial resolution recordings on the surface of the cortex could afford significantly more detailed information regarding microscale dynamics of epileptic electrophysiologic markers otherwise not observable with larger electrode leads. Specifically, we found that interictal discharges take separable, identifiable paths over the cortical surface and that high frequency oscillations can be localized to microscale regions repeatedly over time. Overall, these findings indicate that there may be microdomains of irritative cortex, on the order of 50-100µm, which may be involved in the epileptic network. These events can only be captured with high density surface electrode recordings. The understanding of these dimensions has not been clearly identified in human recordings given lower spatial resolution. The finding of rare microseizure events further illustrates the distinctly local nature of epileptic events. Importantly, these technologies do not penetrate underlying cortex, resulting in an ability to use these devices in eloquent regions with minimal risk to patients, highlighting their translational capability. With ongoing research in the utility of intraoperative recordings for tailoring epilepsy surgery, these data suggest potential benefits of utilizing high spatial resolution recordings both semi-chronically and intraoperatively.

Our current primary limitation of further exploring these phenomena is the limited spatial distribution of the electrode array, which, currently, covers approximately 4×4mm for the circular grid and about 3mm for the bilinear array. Additional limitations include the limited areas of cortical surface that can be sampled, which are dictated by the surgical exposure, as well as current technological limitations in implanting these arrays to allow for semi-chronic recordings. An additional limitation includes the length of the recording, given the need to balance the added operative and anesthetic time for the patient. Nevertheless, even with these limitations, we were able to identify unique features of interictal discharges and high frequency oscillations using superficial microelectrodes otherwise not detectable using macroelectrodes in the acute setting. As intraoperative recordings continue to remain a useful tool for neurosurgeons to excise epileptic foci, our data demonstrate both the feasibility and utility of microelectrode recordings in the operating room. While we recognize that the current dimensions of our microelectrode do not support widespread clinical use given its limited total spatial coverage, we sought to demonstrate, in a translational manner, their initial potential on a feasible scale prior to scaling up.

First, we not only demonstrate that local IID detections can be detected over the cortical surface but also show that IIDs can follow specific, repeated pathways over the cortical surface, as evidenced by the clustered paths in cases where a sufficient number of IIDs were detected. Localized IIDs could correspond to microevents which have been previously reported. In one group of reports, investigators observed these events using penetrating microelectrode semi-chronic recordings in an extraoperative environment (Schevon et al. 2008, 2009, 2010). With the invasive NeuroPort array, which has 400µm electrode spacing, Schevon and colleagues noted the appearance of microdischarges, which were described as epileptiform discharges that could not be seen in adjacent clinical electrodes (Schevon et al. 2008, 2009, 2010). In the 5 patients that were reported in that study, a range of 0 to more than 400 microdischarges were noted in a 5-minute interictal period. In addition, events that occurred on only one microelectrode were documented. Their data were bolstered with further review of the interictal data, which found multiple populations of microdischarges (Schevon et al. 2009). In our study, we similarly found evidence of these microdischarges, however, single channel interictal discharges were not seen, perhaps reflecting the smaller spatial pitch of the PEDOT:PSS devices.

More importantly, by tracking these interictal discharges across the array, we additionally found evidence that interictal discharges may take specific paths across the cortical surface in humans, a finding which may suggest an underlying epileptic network that promotes the progress of interictal discharges across the cortex. Similar findings have been demonstrated previously in animal pharmacologic models (Vanleer et al. 2016), and these IIDs were found to travel at rates similar to those previously reported in animals (primarily in slice physiology), at 29 ± 18 mm/s (Trevelyan et al. 2007).

The study of propagation of IIDs has previously utilized macroelectrode designs and these prior data have been used to understand whether propagation patterns can identify epileptogenic regions (Alarcon et al. 1997; Sabolek et al. 2012; Tomlinson et al. 2016). Specific paths that are taken by interictal discharges are thought to be secondary to specific neuronal responses along the path, coupled with the effect of intervening interneurons (Sabolek et al. 2012; Chizhov et al. 2018). In a clinical context, these propagation patterns have been used to identify specific pathologic regions from which IIDs emanate from, though the ultimate impact on post-surgical outcome is not well defined (Alarcon et al. 1997; Tomlinson et al. 2016). Our method demonstrates that these discharges may in fact be further spatially resolved by microgrids and highlights that a high degree of complexity may exist in how these discharges travel over the cortical surface. Ultimately, these advances may inform future efforts to characterize the underlying epileptic network, particularly at smaller scales, and the efforts to modulate or therapeutically interrupt it.

In addition, prior studies have demonstrated the presence of microseizure events in semi-chronic recordings. Using a subdural electrode grid that included microelectrode contacts with 1 mm pitch, implanted in a semi-chronic setting, Stead and colleagues similarly found examples of seizure-like events that occurred on single microelectrodes, as well as interictal events including interictal epileptiform discharges and high frequency oscillations (HFOs) that occurred on single microelectrode contacts (Stead et al. 2010). Similarly, Schevon and colleagues, using a penetrative microelectrode array with 400µm pitch, demonstrated similar findings with microseizures, which could be found on a single electrode contact (Schevon et al. 2008). While evidence of periodic patterns was noted in our microelectrode data, these did not appear to organize or evolve sufficiently to be deemed seizure events. Nevertheless, we did identify microseizure events as well which were primarily localized to single contacts, with notable voltage spread to neighboring contacts. One benefit of our reduced spatial pitch is that we demonstrate that these events occur over a spatial spread of approximately 100µm.

Our relative paucity of periodic discharges and microseizure events may reflect the relatively short duration of our recordings and the fact that they were performed under anesthesia, either in the form of general anesthesia or sedation. This may have reduced the possibility of capturing seizure events by dampening overall cortical excitability. The administration of provoking medications, such as alfentanil and methohexital, was also insufficient to definitively trigger microseizure events during our microelectrode recordings. Ultimately, as our study is somewhat limited in recording time due to the need to preserve high quality of care for the subjects involved, there are limitations to how many events we could capture over time, which has implications for whether we could observe microseizure events.

By using analyses typically used for neuronal avalanches to uncover repeating spatial and temporal patterns, we demonstrate that HFOs may be detected over the cortical surface in unique, repeatable patterns. As prior clinical studies have suggested, we investigated fast ripples as these phenomena are thought to be more indicative of pathologic activity (Klink et al. 2014). Regarding high frequency oscillations, Schevon and colleagues additionally reported the appearance of HFOs on their penetrative microelectrode array and noted that the majority of their detections only involved a single channel (Schevon et al. 2009). Interestingly, they found that microdischarges and HFOs tended to have some degree of independence between electrode sites. While our data demonstrated similar examples of HFOs, we expand on these prior findings by demonstrating that HFOs can be detected on non-penetrative microelectrodes and that specific areas of the cortical surface, as sensed by our PEDOT:PSS microelectrode devices, tended to fire in concert, perhaps demonstrating how specific regions of the cortical surface may be more epileptogenic. These results again highlight the complex underlying cortical architecture that can lead to seizures.

In general, macroelectrodes have been used to identify HFOs in seizure onset zones. Overlap between HFOs and seizure onset zones has been demonstrated, and HFOs have been considered to be favorable markers for the surgical targeting (Zijlmans et al. 2011; Korzeniewska et al. 2014; Schönberger et al. 2019). However, one prospective trial recently demonstrated that using HFOs to guide surgical resection to predict ultimate seizure outcome is challenging given conflicting clinical data regarding the benefit of using HFOs intraoperatively, highlighting potential network phenomena that link HFOs (Jacobs et al. 2018). Intraoperatively, HFOs, and particularly fast ripples, have been identified as an important predictor of post-operative outcome, with the presence of residual fast ripples in post-resection recordings potentially predicting post-operative outcome (Klink et al. 2014; Hussain et al. 2017; Klooster et al. 2017). In addition, there has been concern that due to fast ripples representing small areas of cortex, undersampling can occur with current clinical intraoperative macroelectrode recordings which are on the scale of millimeters (Klooster et al. 2017). Our study builds on this prior work by demonstrating specific spatiotemporal patterns can occur in microdomains with regard to HFOs. We demonstrate that specific regions of HFOs can fire in specific patterns, which supports the idea of an underlying epileptic network. Moreover, the high spatial resolution of our PEDOT:PSS microgrids shows that HFOs can be detected over small regions of the cortical surface, in which single-channel events could be seen. Our work similarly supports prior microelectrode-based HFO findings that demonstrated that focal HFOs can be identified in regions < 1 mm^3^ (Worrell et al. 2008).

Our work with multi-unit activity demonstrated a relationship with HFOs, but not with IIDs. Some reasons for this may include limitations to which surface microelectrode are able to sample neural activity at depth, as well as the limited number of MUA clusters that were identified. It may additionally suggest that the HFO activity detected originates at a more local level, as compared to the IID activity, which may require firing of a larger network of neurons. Prior data has demonstrated that single units may have different firing patterns during the temporal course of an IID (Keller et al. 2010). In the context of human mesial temporal epilepsy, Alvarado-Rojas and colleagues demonstrated that approximately 40% of single units demonstrated a change in firing (Alvarado-Rojas et al. 2013). In addition, they hypothesized that heterogeneous, sparse populations could be involved in the generation of IIDs (Alvarado-Rojas et al. 2013). These data have been additionally built upon by other work that has similarly demonstrated changes in firing rates and patterns of single units in the setting of IIDs (Keller et al. 2010; Alarcón et al. 2012). Given that a substantial fraction of neurons may not be involved in IIDs, the limited clusters of MUA in our study may preclude our ability to find a definitive relationship. With regard to single unit activity and ripples in humans, prior groups had demonstrated that single units in the hippocampus demonstrate changes in firing during ripples (Quyen et al. 2008), a relationship that extends similarly to fast ripples (Köhling and Staley 2011; Jiruska et al. 2017). Our results may therefore be explained by the microelectrode detecting both fast ripples and MUA activity at a local, superficial level.

Finally, we demonstrate that using PEDOT:PSS microelectrodes in an acute intraoperative setting is feasible in a large number of settings and patients. The results of being able to detect HFO active zones or IID paths using these microscale detection approaches could have wide-reaching implications and offer a more directed set of tools to identify irritable tissue. Future work will focus on increasing the number of electrodes on the array to expand spatial coverage, which has been the direction of the field of advanced microelectrode designs (Chiang et al. 2020). In addition, an exciting direction would be to enable within-operating room visualizations of these localized HFO regions of interest and IID waves. Indeed, by highlighting that such technologies may be able to assist in neurosurgical procedures in the future, we may be able to better understand the underlying seizure network as well as targeting irritable cortical tissue.

## Data Availability

The data that support the findings of this study are available from the senior author, upon reasonable request.

## Acknowledgements and Sources of Funding

We would like to thank Yangling Chou, Erica Johnson, and Gavin Belok for their assistance in data collection. We also thank Melissa Murphy and Aaron Tripp and other members of the intraoperative monitoring teams at BWH and MGH for their assistance with intraoperative clinical recordings. Finally, we would like to recognize the patients for their invaluable participation in the study.

This research was sponsored by the U.S. Army Research Office and Defense Advanced Research Projects Agency under Cooperative Agreement Number W911NF-14-2-0045. In addition, other support included ECOR and K24-NS088568 to SSC; NSF-CAREER award #1351980, NSF CMMI award #1728497, and NIH DP2-EB029757 to SAD. The views and conclusions contained in this document are those of the authors and do not represent the official policies, either expressed or implied, of the funding sources.

## Ethical Publication Statement

We confirm that we have read the Journal’s position on issues involved in ethical publication and affirm that this report is consistent with those guidelines.

